# Comparing the efficacy of Ventriculoperitoneal Shunts with Lumboperitoneal Shunts in the treatment of Idiopathic Normal Pressure Hydrocephalus: A Systematic Review and Meta-Analysis

**DOI:** 10.64898/2026.01.07.26343481

**Authors:** Arya Abaee, Owen David Kelly, Lewis Thorne

## Abstract

**Introduction:** Ventriculoperitoneal (VP) and Lumboperitoneal (LP) shunts are the most common treatments for Idiopathic Normal Pressure Hydrocephalus (iNPH). Shunt procedure choice is generally based on surgeon on preference rather than evidence. We performed a systematic review and meta-analysis to address this gap for evidence-based shunt selection in iNPH treatment.

**Methods:** Publications on post-operative outcomes for LP and VP shunts in iNPH were identified in MEDLINE and EMBASE. Papers were selected based on pre-specified inclusion and exclusion criteria and meta-analysis was conducted for outcome measures after shunt procedure.

**Results:** 17 papers were included. LP Shunt patients showed greater cognitive improvement with an average increase of 2.00 points (95% CI: 1.08; 2.93, p < 0.0001) on their MMSE score post-operatively compared to VP shunt patients who improved on average by 1.30 points (95% CI: 0.81; 1.79, p < 0.0001). The LP group had considerable heterogeneity (I^2^ = 66.42%, p = 0.0003) whereas the VP shunt group had minimal heterogeneity (I^2^ = 0.00%, p = 0.8447) reflecting more uniformity across its included studies. For overall symptomatic improvement measured by the iNPHGS, VP shunts patients demonstrated a larger reduction in overall symptom scores with an average decrease of 2.91 points (95% CI: -3.78; -2.05, p < 0.0001) but with a high heterogeneity (I^2^ = 79.12%, p = 0.0012) compared to LP shunt patients with an average reduction of 1.91 points (95% CI: -2.31; -1.51, p < 0.0001) with no detected heterogeneity (I^2^ = 0.00%, p = 0.8454).

**Conclusions:** Our findings demonstrate that LP and VP shunts show differing patterns of improvement across the cognitive domain and the broader iNPH triad, with LP shunting showing greater cognitive improvement and VP shunting showing greater overall symptomatic improvement. These differences represent a signal warranting further investigation, specifically whether symptom profiles should inform shunt selection.

## Introduction

Shunt surgery is the current gold standard treatment and diagnosis for idiopathic normal pressure hydrocephalus (iNPH). Shunting aims to move Cerebrospinal Fluid (CSF) to another anatomical cavity where it can be effectively reabsorbed (Williams and Malm, 2016). There is heterogeneity in options available to neurosurgeons when considering shunting, both in the type of procedure (Yamada *et al*., 2022) and shunt valve mechanism (Miyake, 2016). Currently there is no clear standardisation regarding this choice, with many of these decisions left to the patient and surgeon’s personal preference (Turner, 1995), (Subramanian *et al*., 2018) with potential variations in patient outcomes (Nakajima *et al*., 2021).

We classify shunts by the anatomical compartments they connect, e.g. Ventriculoperitoneal (VP), Lumboperitoneal (LP), and Ventriculoarterial (VA) (Graham *et al*., 1982). For the purposes of this paper, we’ve concentrated on these two most common shunt types which are VP and LP.

VP shunts are the most commonly used treatment internationally for iNPH, with usage rate found by a national study in the US looking at 60,000 patients to be 72% (Alvi *et al*., 2021) (Nakajima *et al*., 2021). VP shunts consist of a proximal catheter, used to drain cranial CSF, generally placed in the right lateral ventricle; a distal catheter with its end in the peritoneal cavity where the CSF can be reabsorbed (Williams and Malm, 2016), and a valve connecting the two, regulating the intracranial pressure above which CSF is drained (Sahuquillo *et al*., 2021). LP shunts, whilst essentially working with the same mechanism and parts, has the proximal catheter placed in the subarachnoid space of the spinal cord in the lumbar region (Medtronic, 2023), rather than in the cranium and thus drain lumbar CSF. Overall, they are less commonly used (Alvi *et al*., 2021), but regional fluctuations exist: in Japan LP shunts are more common than VP shunts (Nakajima *et al*., 2021). As catheter designs are relatively uniform across manufacturers and companies, their effect on outcomes is likely negligible (Sahuquillo *et al*., 2021).

Further complicating surgical decision-making is valve choice for the shunt used. Shunt valves have multiple variations - they can either be fixed or programmable and can come with an anti-siphon or gravity-compensating component (Ahmed *et al*., 2023). Fixed valves maintain a constant opening pressure that cannot be adjusted post-implantation (Rinaldo *et al*., 2020), whereas programmable valves allow for non-invasive adjustments to the opening pressure over time (Serarslan *et al*., 2017), and thus can non-invasively solve drainage-related issues (Scholz *et al*., 2018). This factor has led programmable values to be effective in managing iNPH by enabling fine-tuned control over cerebrospinal fluid (CSF) drainage (Delwel *et al*., 2013), but are not as cost-effective as their fixed counterparts (Agarwal *et al*., 2019). Admittedly, there remains currently no strong evidence-base to support use of a specific model of shunt valve over another (Bergsneider *et al*., 2005).

VP and LP shunts have potentially different physical properties but there are also differences in lumbar and cranial CSF. A paper by Rostgaard et al. in 2023 compared cranial and lumbar CSF in an iNPH patient population. In a proteomic analysis, they found 22 proteins present in ventricular CSF that were not present in lumbar CSF. They also reported differing abundances of 216 common proteins between cranial and lumbar CSF (Rostgaard *et al*., 2023). Pairing this information with findings of glymphatic clearance impairment in iNPH patients (Ringstad, Vatnehol and Eide, 2017) leads us to speculate on the practical differences between VP and LP shunting on proteomic collections in the two distinct regions prior to, and post, shunting.

The inherent heterogeneity of iNPH presentation and vast number of treatment options has led to a significant gap in the literature regarding comparing shunt placement choices. The authors of this study were able to find only one other meta-analysis attempting to compare VP and LP shunts in iNPH (Giordan *et al*., 2019), which concentrated primarily on responsiveness and ignored other critical outcome measures that typically influence surgical decision making such as: complication rate (Kobayashi *et al*., 2022), revision rate (Hung *et al*., 2016) and quantitative measures of improvement (Van Swieten *et al*., 1988) (Kubo *et al*., 2008).

It is our personal experience that many surgeons choose their shunt, *“in much the same way they buy a car. To varying degrees, they use style, past experience, brand loyalty, advertising, comfort, training, and a little science to choose a combination of hardware and surgical procedure. The choice becomes very emotional to many…”* (Turner, 1995). Many studies have examined the safety and efficacy of a single shunt type, be it VP or LP, but the choice of implant is largely due to individual surgeons’ preference. A systematic review and meta-analysis of the current literature regarding VP and LP shunts in iNPH could potentially provide a framework to formulate a personalised gold standard selection method for the most appropriate shunt type for any one individual on the basis of preoperative evaluations.

To quantify the effect of VP and LP shunting, we selected the Mini Mental State Examination (MMSE) and the Idiopathic Normal Pressure Hydrocephalus Grading Scale (iNPHGS) to compare shunt effectiveness, as these are the most commonly reported outcome measures in iNPH literature (Nakajima *et al*., 2021).

The MMSE is a general cognitive screening tool designed for broad use across a spectrum of conditions including iNPH, Dementia and head injury to name a few. It is the most widely used test for standardised cognitive assessment as a result (Gallegos *et al*., 2022). It tests six main cognitive domains of orientation, registration, attention, recall, language and visuospatial construction; as cognition improves, patient’s scoring increases up to a maximum of 30 points. Within iNPH, higher preoperative MMSE scoring correlates with greater improvement in working memory and language post operatively (Xiao *et al*., 2022). Therefore, a quantitative improvement is based on the difference between pre- and post-operative scores.

However, as iNPH is characterised by a triad of symptoms where cognition is only one domain, the MMSE alone cannot capture the full clinical picture. The iNPHGS bridges this gap and correlates strongly with other useful diagnostic tests (Kubo *et al*., 2008). It scores the separate domains of the iNPH triad with higher scores in each indicating greater symptom severity. By comparing both the iNPHGS and MMSE, we are able to assess a range of treatment responses. Our meta-analysis employs a random-effects model to account for heterogeneity in patient populations (Viechtbauer, 2010).

## Methods

### Search Strategy

A comprehensive search of both MEDLINE and EMBASE databases were used to identify relevant articles. The search strategy was created and then reviewed by a clinical librarian with the following search being employed:

1. ("Ventriculoperitoneal Shunt" or "Ventriculoperitoneal Shunts" or "Ventricular Shunt*" or "VP Shunt*" or "VPS" or "Ventriculoperitone*").af.
2. ("Lumboperitoneal Shunt*" or "Lumbar Shunt*" or "LP Shunt*" or "Lumboperitone*" or "LPS").af.
3. 1 OR 2
4. ("Normal Pressure Hydrocephalus" or "Idiopathic normal-pressure hydrocephalus" or "iNPH").af.
5. 3 AND 4

## Inclusion and Exclusion Criteria

Available articles were chosen if they fulfilled the following inclusion criteria:

- Articles involving Ventriculoperitoneal shunts and/or Lumboperitoneal Shunts
- Articles must report at least one outcome measure as stated in the Methodology section of this paper i.e. iNPHGS score, MMSE
- Articles must report the pre and post intervention average means for the populations with standard deviations or provide patient specific data
- Articles must report their methodology for patient selection and must report both clinical diagnosis and at least one form of diagnostic testing as selection criteria i.e. Imaging etc.
- Articles must follow up patients at least 1-year post shunt surgery
- Articles must report their methodology for statistical analysis
- Articles must be available in the English Language
- Articles must be published after 2009

Available articles were rejected based on the following exclusion criteria:

- Articles involving Ventriculoarterial shunts only
- Systematic reviews, narrative reviews, scoping reviews, meta-analyses, non-peer reviewed studies, conference abstracts, letters or commentaries
- Case reports with less than 10 patients

## Screening and Data extraction

All available studies were codified into a CSV file and duplicates were removed. Using Excel, the articles went through a full title, abstract and article review by two separate fully blinded

reviewers (A.A and O.K). Once extraction was finished, any disagreements between the two reviewers were settled through discussion. Data extraction then occurred separately with two data extraction tables being created after the articles full text was examined.

## Statistical Analysis

Pre- and post-operative scores for the MMSE and the iNPHGS were extracted from the included articles and mean change scores, their standard deviations and confidence intervals calculated for each paper. The “metafor” package (Viechtbauer, 2010) in R studio 12.1 was utilised with the DerSimonian and Laird random-effects model to carry out the statistical analysis (DerSimonian and Kacker, 2007). Statistical heterogeneity was quantified using the I^2^ statistic. GraphPad Prism was employed to generate forest plots (GraphPad Software, 2024).

## Results

### Search Results

1717 papers were identified in the search (Fig. 1). Once duplicates were removed 1362 articles were screened, of which 16 adhered to the inclusion and exclusion criteria and were included in the final meta-analysis: 9 articles had LP shunting as the exclusive treatment and 8 articles had VP shunting as the exclusive treatment. These 16 articles encompassed 803 patients; 302 patients received LP shunts and 501 patients received VP shunts. One article (Lukkarinen et al., 2022) included cohorts from two separate countries and was treated as two separate articles in the analysis.

**Fig 1.**
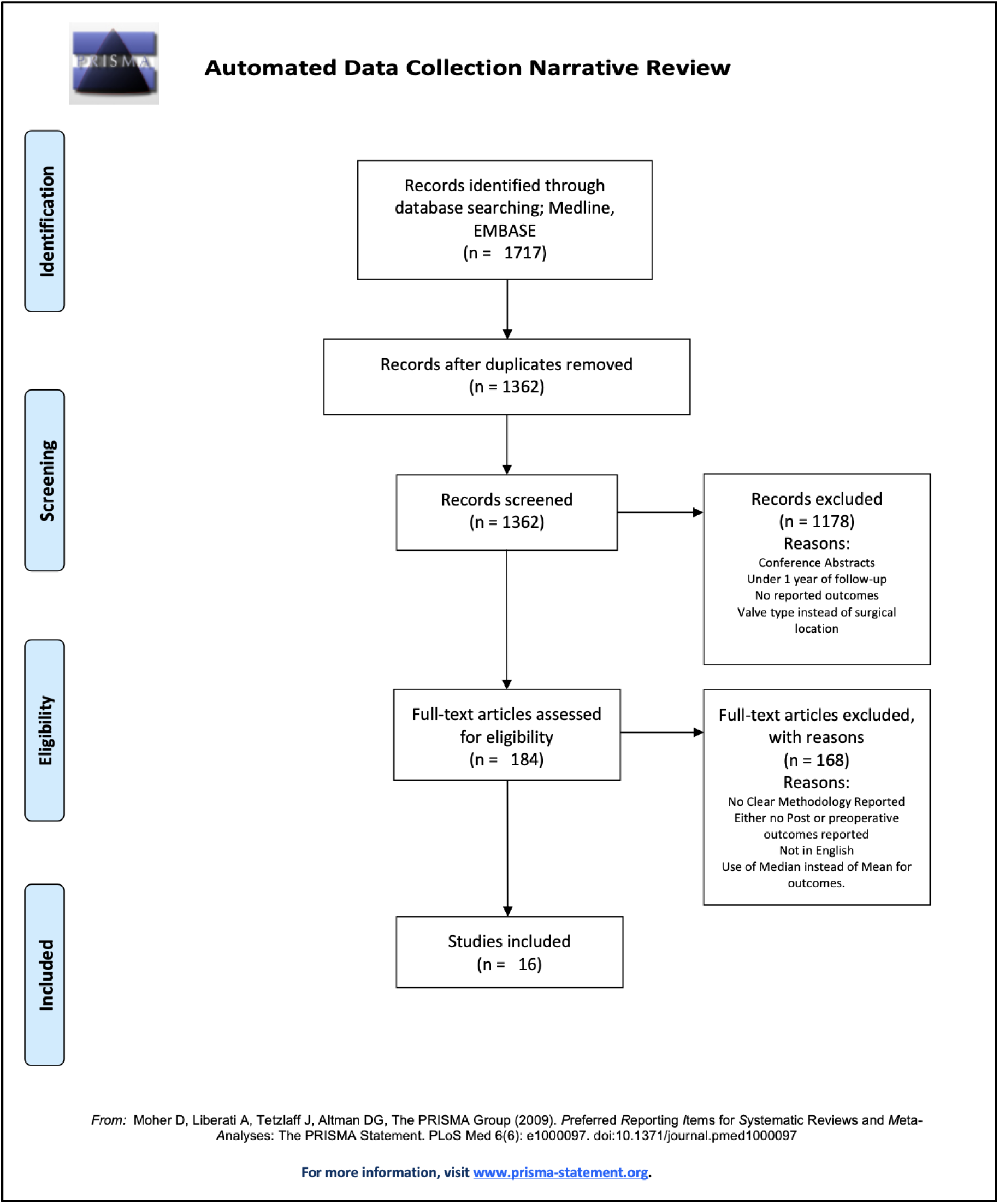
**PRISMA Flow diagram of article screening process (Page *et al*., 2021)**. A total of 16 articles were collated from the literature that adhered to inclusion and exclusion criteria.

### MMSE

Seven articles reported pre- and post-operative scores for the MMSE with an intervention of LP shunting (Fig. 2; Table 1.). The pooled mean change after LP shunting was an increase of 2.00 points (CI:1.08, 2.93 P<0.0001(***)) in the MMSE score. There was substantial heterogeneity noted within the group by the I^2^ statistic of 66.42% (P=0.003*).

**Fig 2.**
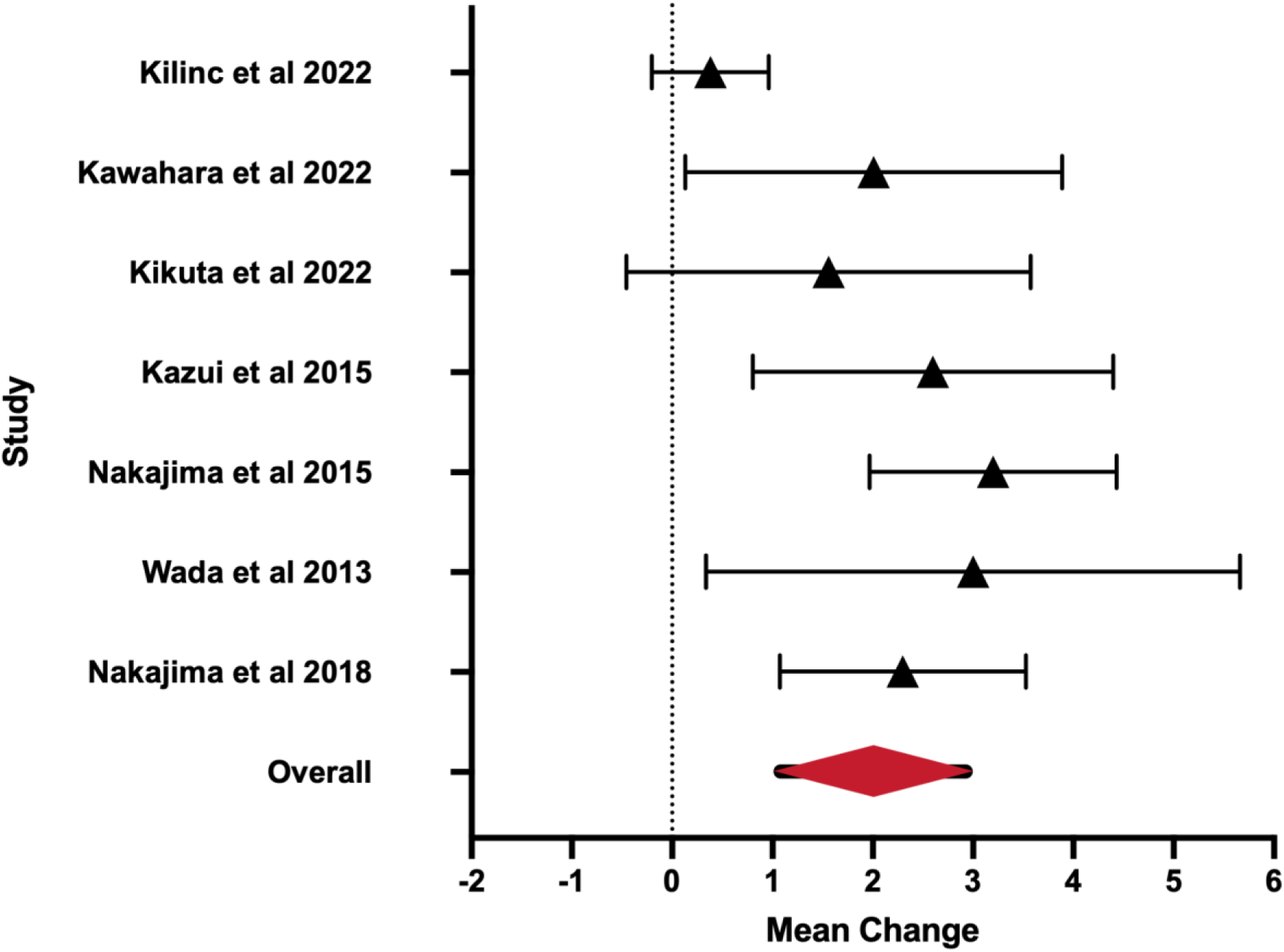
Forest Plot for MMSE - LP Shunt. The error bars represent the 95% confidence intervals with the mean change represented by the red triangle. The dashed line represents a mean change of 0.

**Table 1.** Mean change scores for MMSE scores - LP shunt. . N = number of study participants; CI = confidence interval.

| **Study** | **N** | **Mean Change** | **CI - Lower** | **CI - Upper** |
| --- | --- | --- | --- | --- |
| Kilinc et al 2022 | 42 | 0.38 | -0.20 | 0.96 |
| Kawahara et al 2022 | 45 | 2.01 | 0.13 | 3.89 |
| Kikuta et al 2022 | 09 | 1.56 | -0.46 | 3.58 |
| Kazui et al 2015 | 40 | 2.60 | 0.80 | 4.40 |
| Nakajima et al 2015 | 51 | 3.20 | 1.97 | 4.43 |
| Wada et al 2013 | 12 | 3.00 | 0.34 | 5.66 |
| Nakajima et al 2018 | 64 | 2.30 | 1.07 | 3.53 |
| **Overall** | **263** | **2.00** | **1.08** | **2.93** |

Four articles reported pre- and post-operative scores for the MMSE with the intervention of VP shunting (Fig. 3; Table 2). The paper by Lukkarinen et al 2022 included two separate cohorts administered by two separate centres and was thus treated as separate articles. The pooled mean change after VP shunting was an increase of 1.30 points (CI:0.81, 1.79 P<0.0001(***)) in the MMSE score. There was no heterogeneity noted within the group by the I^2^ statistic of 0.00%, though this was not a significant finding (P=0.8447).

**Fig 3.**
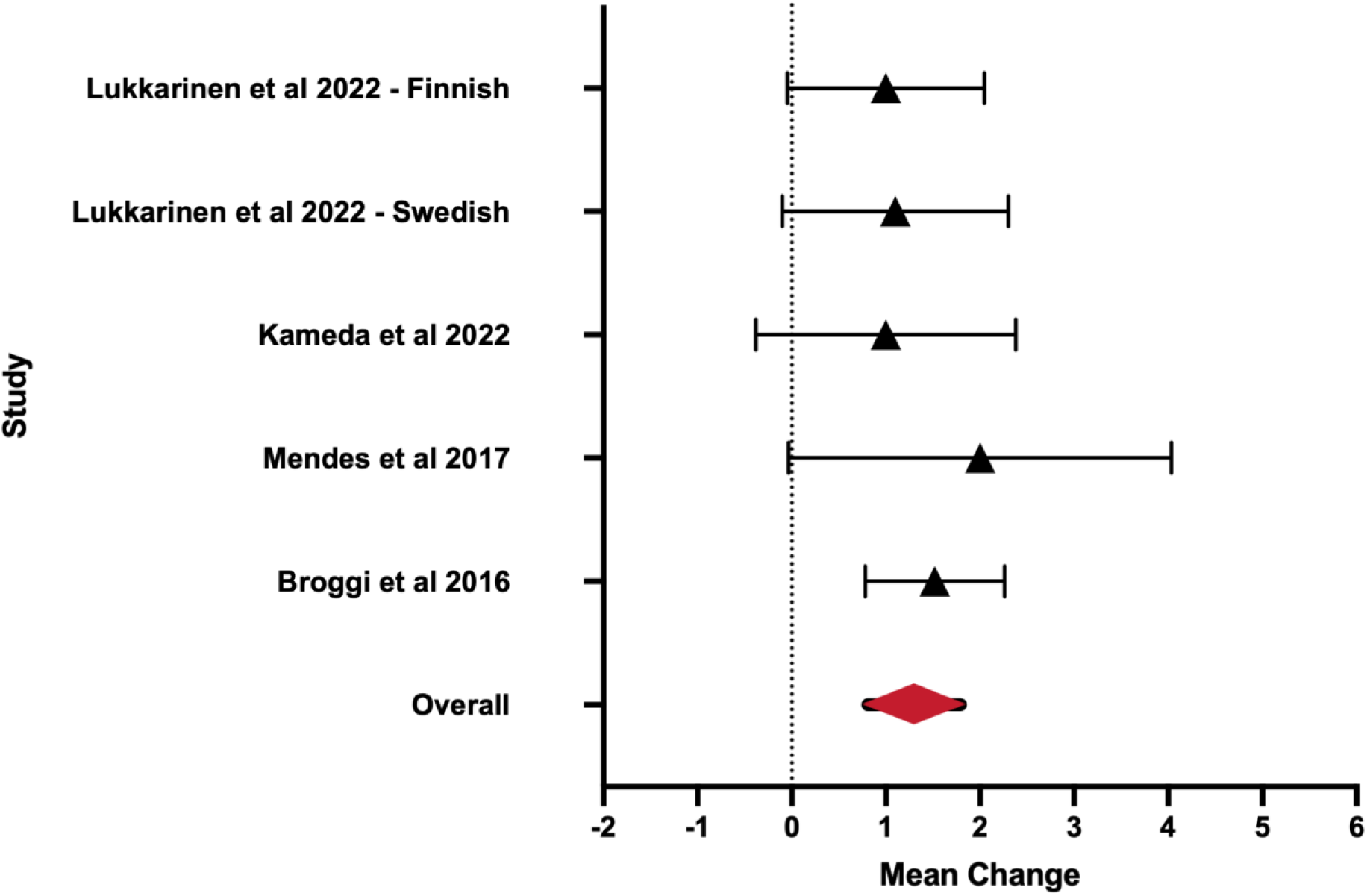
**Forest Plot for MMSE - VP Shunt**. The error bars represent the 95% confidence intervals with the mean change represented by the red triangle. The dashed line represents a mean change of 0.

**Table 2.**
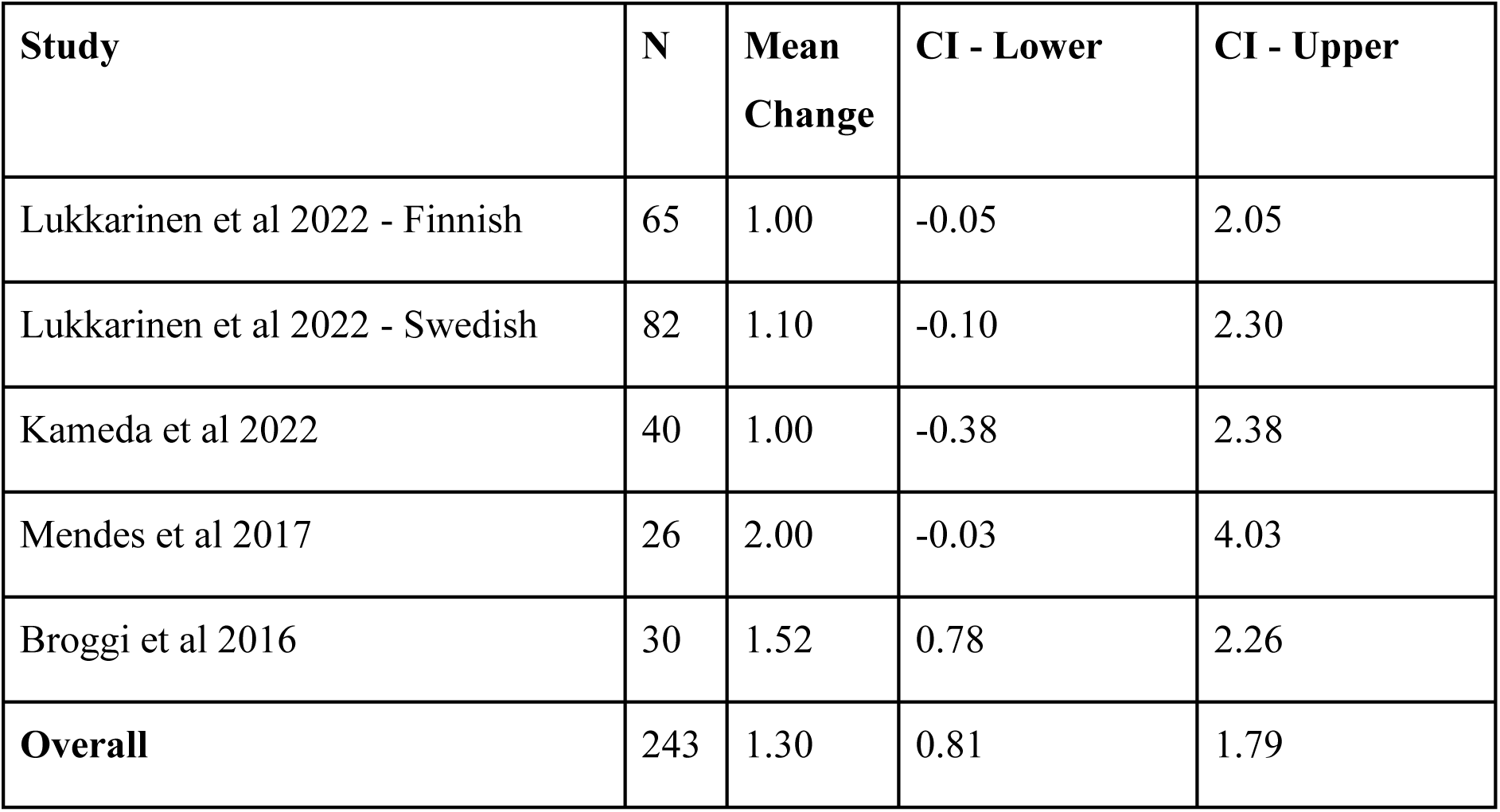
Mean Change scores for MMSE scores - VP Shunt. N = number of study participants; CI = confidence interval.

### iNPHGS

Three articles reported the pre- and post-operative scores for the iNPHGS with the intervention of LP shunting (Fig. 4; Table 3). The pooled mean change score after LP shunting showed a reduction of 1.91 points in the iNPHGS (CI:-2.31, -1.51 P<0.0001(***)). There was no heterogeneity noted within the group by the I^2^ statistic of 0.00%, though this was not a significant finding (P=0.8454). The average change score of all of the articles were significant as no confidence interval crosses 0.

**Fig 4.**
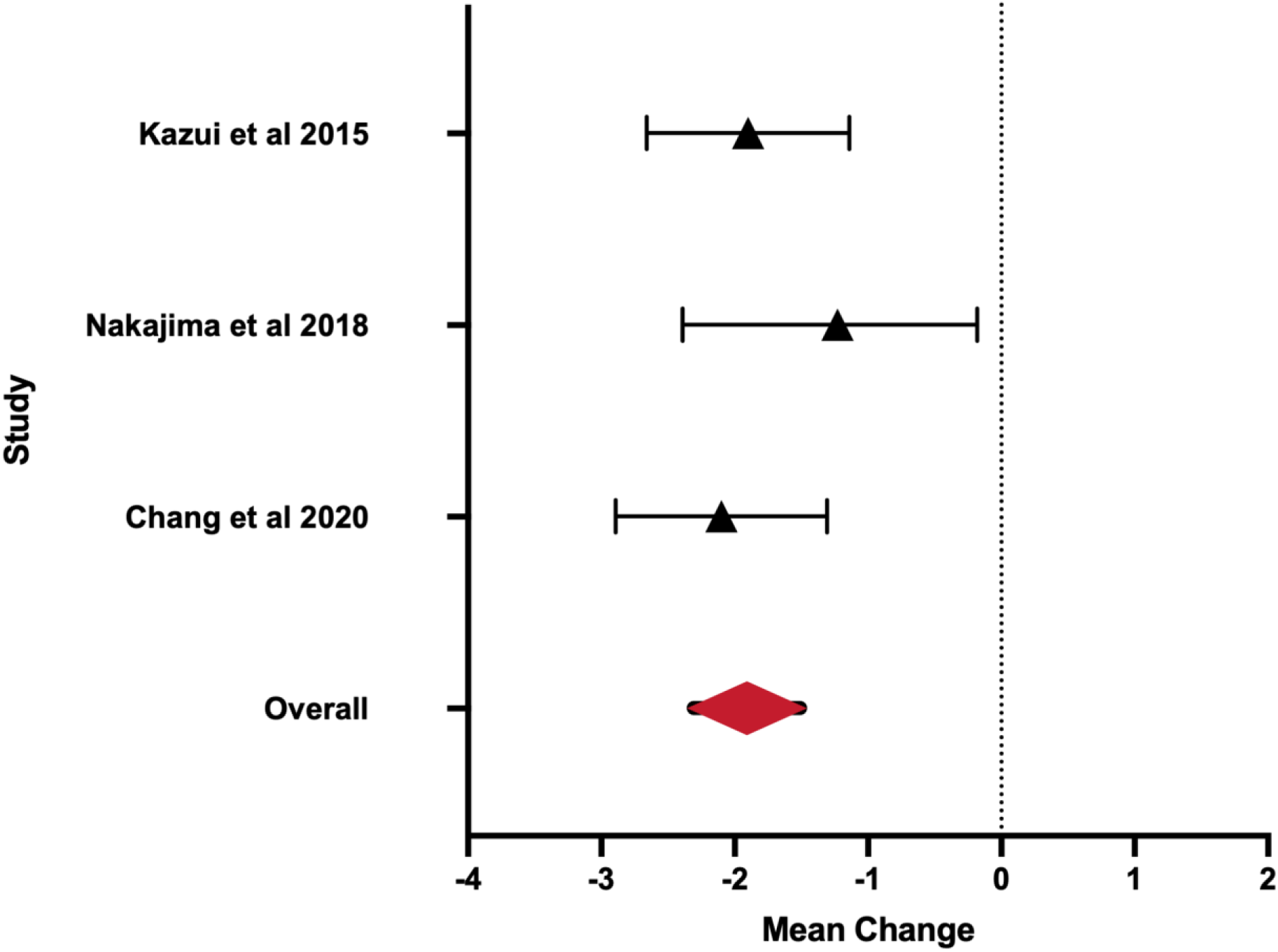
Forest Plot for iNPHGS - LP Shunting. The error bars represent the 95% confidence intervals with the mean change represented by the red triangle. The dashed line represents a mean change of 0.

**Table 3.**
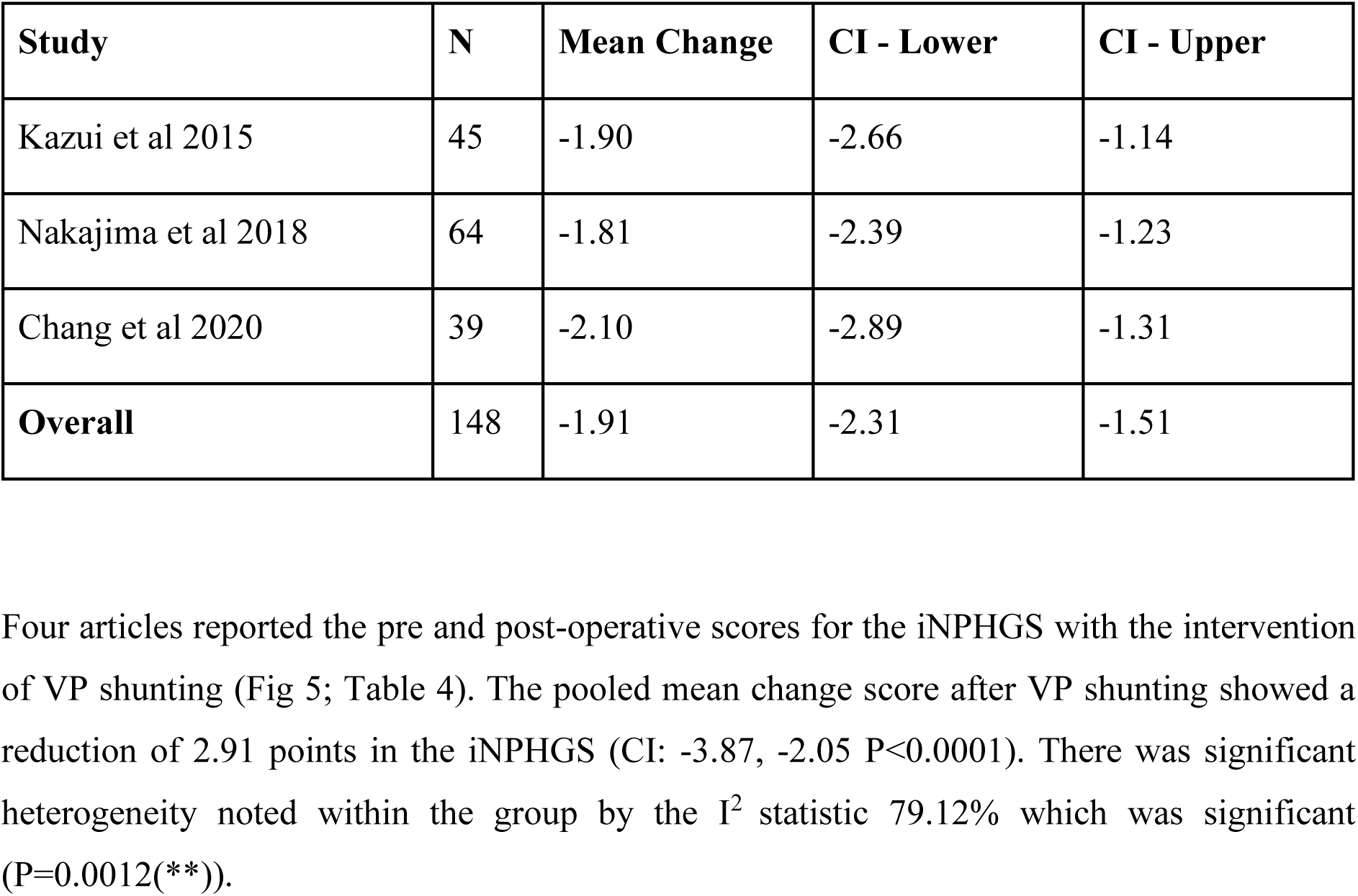
Mean Change Scores for iNPHGS scores - LP Shunting. N = number of study participants; CI = confidence interval.

Four articles reported the pre and post-operative scores for the iNPHGS with the intervention of VP shunting (Fig 5; Table 4). The pooled mean change score after VP shunting showed a reduction of 2.91 points in the iNPHGS (CI: -3.87, -2.05 P<0.0001). There was significant heterogeneity noted within the group by the I^2^ statistic 79.12% which was significant (P=0.0012(**)).

**Fig 5.**
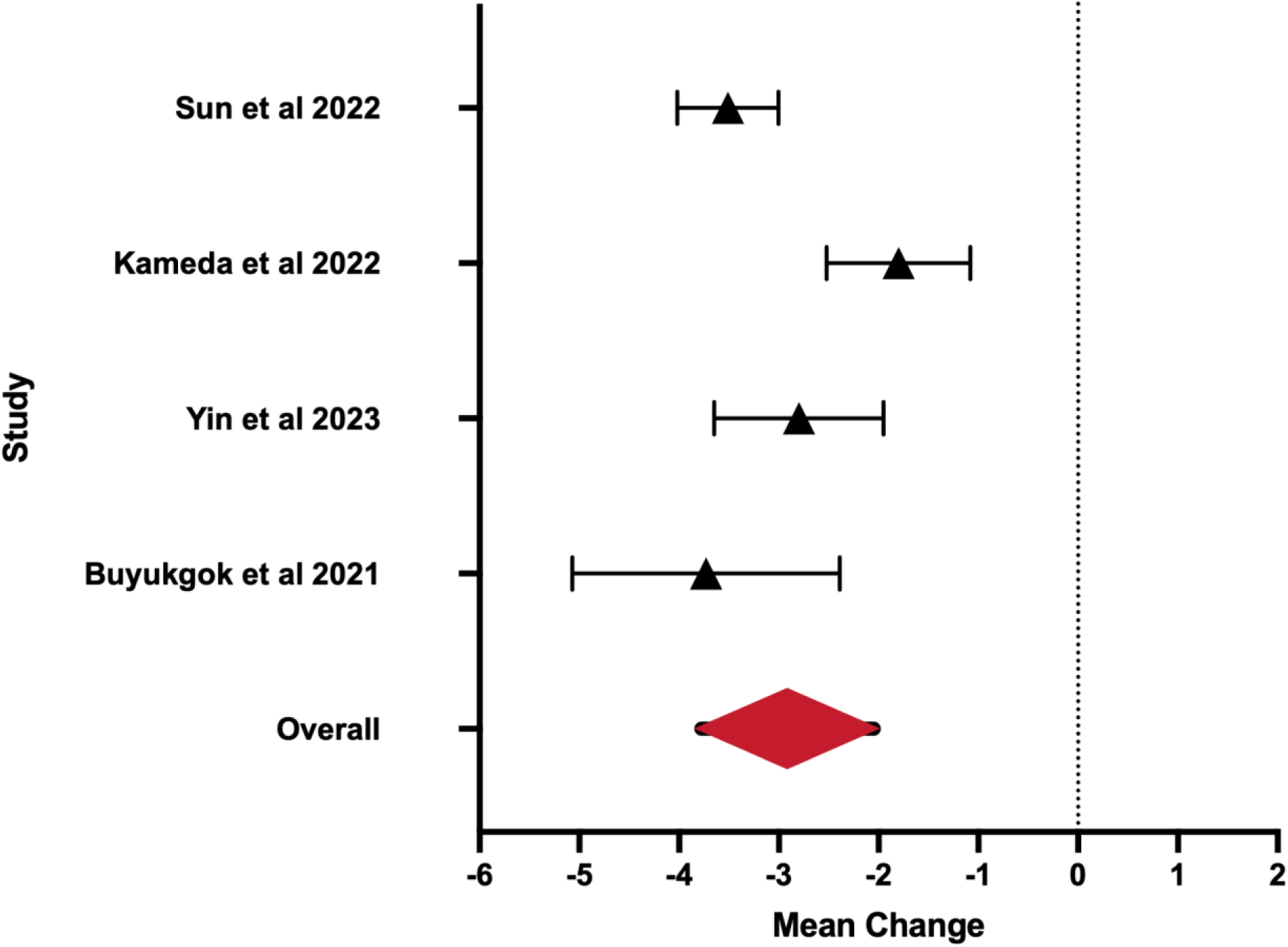
Forest Plot for iNPHGS - VP Shunting. The error bars represent the 95% confidence intervals with the mean change represented by the red triangle. The dashed line represents a mean change of 0.

**Table 4.**
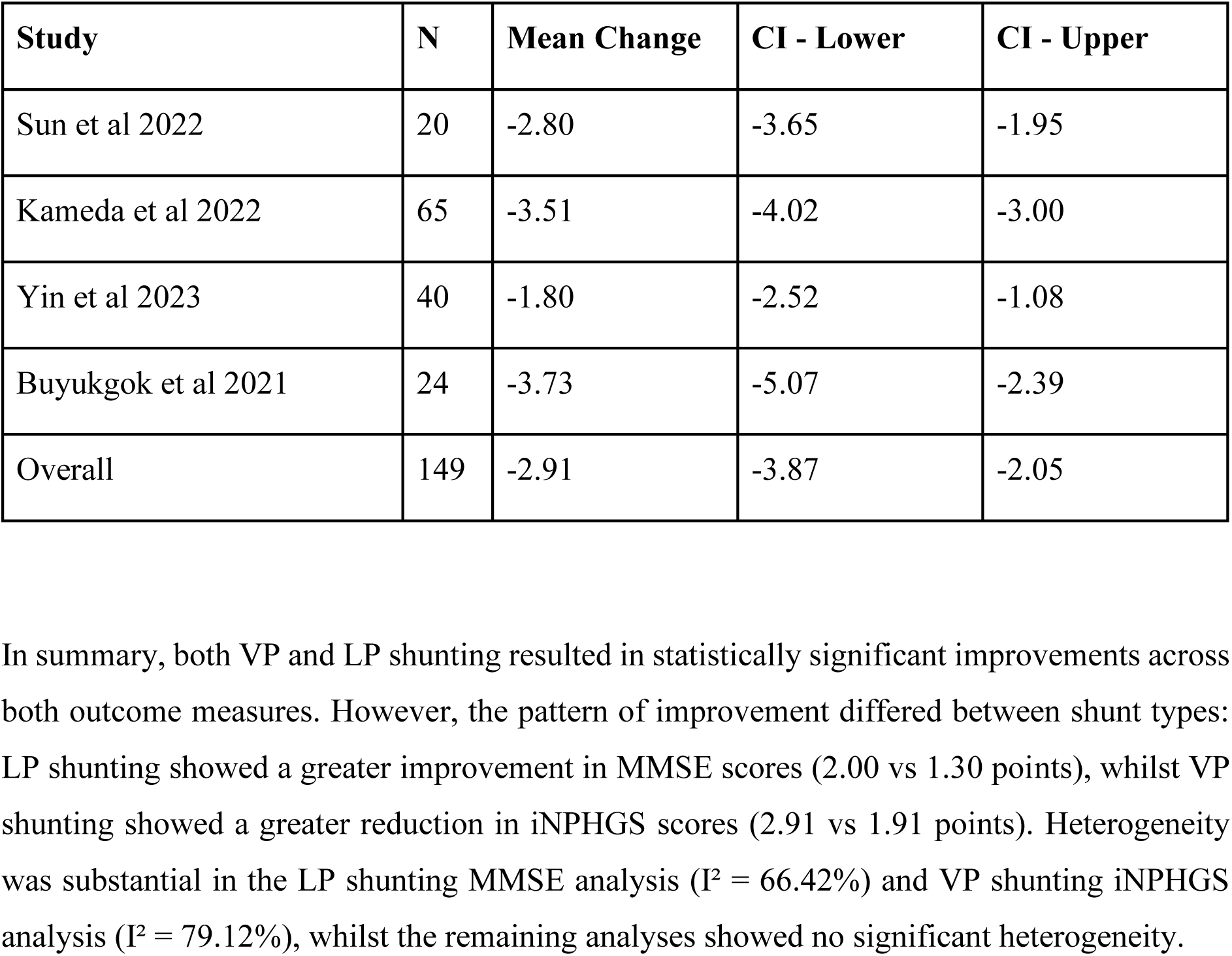
Mean Change Scores for iNPHGS scores - VP Shunting. . N = number of study participants; CI = confidence interval.

In summary, both VP and LP shunting resulted in statistically significant improvements across both outcome measures. However, the pattern of improvement differed between shunt types: LP shunting showed a greater improvement in MMSE scores (2.00 vs 1.30 points), whilst VP shunting showed a greater reduction in iNPHGS scores (2.91 vs 1.91 points). Heterogeneity was substantial in the LP shunting MMSE analysis (I² = 66.42%) and VP shunting iNPHGS analysis (I² = 79.12%), whilst the remaining analyses showed no significant heterogeneity.

## Discussion

This meta-analysis aimed to address the comparative ambiguity around the efficacy of ventriculoperitoneal shunting vs lumboperitoneal shunting in improving symptoms in patients with iNPH. The MMSE was selected to assess cognitive change specifically, whilst the iNPHGS captures the overall symptom profile across the triad (Kubo *et al*., 2008). The differences observed between VP and LP shunting across these measures represents a signal warranting further investigation, specifically whether symptom profiles could inform shunt selection choice. Though given the retrospective nature of the data in this meta-analysis, these findings should be viewed as a foundation for future research.

It is important to note the assumptions underlying our interpretation of these results. The iNPHGS provides a total score across all three symptom domains, meaning we cannot determine which specific symptom domain is driving the observed improvement. We have therefore assumed an average reduction across domains when interpreting the iNPHGS change scores. However, the fact that we observe different patterns of improvement between VP and LP shunting in both the MMSE and iNPHGS suggests that the non-cognitive domains may also respond differently depending on shunt type, though this cannot be confirmed without domain-specific data which does not seem to be readily available across the literature.

Generally, literature comparing LP and VP shunting in iNPH consist of comparative single centre studies or some multi-centre trials (Xie *et al*., 2021) (Moran *et al*., 2016). One other meta-analysis has been identified as mentioned above (Giordan *et al*., 2019). However, meta-analyses comparing LP and VP shunting in other forms of hydrocephalus have occured (Ho *et al*., 2023) (Andreão *et al*., 2024). These studies report that LP and VP shunts are comparable in their effects with some differences in the complication rate and revision rate. Studies in this field typically employ odds ratios to determine whether patients exhibit a response to an intervention, but fail to quantify its magnitude. As a result, the true extent of improvement when comparing two interventions is obscured. This is a critical factor in surgical decision making in iNPH where a broad spectrum of cognition, gait and urinary symptoms intermingle.

By examining the effect size, our results demonstrate that there is a clear difference between LP and VP shunting when subdomains of iNPH are considered. Our results show that LP shunting results in a greater improvement in the MMSE score whilst VP shunting results in a greater improvement in the iNPHGS score. Although it would be premature to conclude that LP shunting should be used for patients with more serious cognitive symptoms while VP should be deployed in patients to address the overall symptom profile, these results indicate that shunt choice may have differing benefits across iNPH’s symptomatic domains.

It remains unclear whether these results would translate to what might be a meaningful clinical difference for a patient and more experimental more real-world data will be required. But to speculate further: symptom profiles of patients might be considered when deciding on the primary method of shunting. In fact, this may require a shift in the “either-or” paradigm that currently exists within the literature, where the main question is whether VP or LP shunting is better, to a more nuanced approach where patient symptom profiles lead to shunt choice.

It is important to note several limitations within this meta-analysis. Firstly, the high level of heterogeneity within both the patient population i.e., in age and comorbidities, as well as variability within research protocols between centres, making comparison and pooling of data more tenuous. An example of this is the follow-up time frame; this was chosen based on our original pilot study of papers to allow us to encompass the highest number of studies whilst reducing the effect of concurrent neurodegenerative diseases such as Alzheimer’s and Parkinson’s Disease, which are common in patients with iNPH (Hu *et al*., 2024). Secondly, the use of single centre studies as highlighted by Azad et al., 2020 (Azad *et al*., 2020) suggests that they emphasise the impact of institutional preferences i.e. a centre that more often prefers to insert VP shunts may show better results for VP shunt patients which once again impacts our meta-analysis. Additionally, reporting outcomes cannot be blinded in all cases as the centre is aware of what operation it is performing and often the surgeon themselves may also perform the follow up testing, thus inserting a potential for bias. We also did not examine complication or revision rates directly, because definitions and follow-up durations varied so widely between studies that a pooled estimate would be unreliable.

A network meta-analysis could overcome these limitations. By allowing both direct and indirect comparisons, this methodology could incorporate single-arm studies, link them through shared comparators, and account for multiple interventions. This expanded approach would capture a wider range of clinical data while potentially mitigating biases stemming from institutional preferences. As a result, it may yield more robust and generalisable findings regarding the comparative efficacy of LP and VP shunting in iNPH.

In summary, our meta-analysis found that there is a clear difference in the efficacy of LP and VP shunting. Importantly, each showed differing patterns of improvement across the cognitive domain and the broader iNPH triad. Further research to determine the patient profile symptom clusters and stratify our intervention by cluster may lead to a more robust surgical decision making for shunt treatment of iNPH.

## Data Availability

All data produced in the present study are available upon reasonable request to the authors

## References

1. Agarwal, N., Kashkoush, A., McDowell, M. M., Lariviere, W. R., Ismail, N. and Friedlander, R. M. (2019). ‘Comparative durability and costs analysis of ventricular shunts’. Journal of Neurosurgery, 130 (4), pp. 1252–1259. doi: 10.3171/2017.11.JNS172212.

2. Ahmed, M., Naseer, H., Farhan, M., Arshad, M. and Ahmad, A. (2023). ‘Fixed versus Adjustable differential pressure valves in case of idiopathic normal pressure hydrocephalus treated with ventriculoperitoneal shunt. A systematic review and meta-analysis of proportion’. Clinical Neurology and Neurosurgery, 230, p. 107754. doi: 10.1016/j.clineuro.2023.107754.

3. Alvi, M. A., Brown, D., Yolcu, Y., Zreik, J., Javeed, S., Bydon, M., Cutsforth-Gregory, J. K., Graff-Radford, J., Jones, D. T., Graff-Radford, N. R., Cogswell, P. M. and Elder, B. D. (2021). ‘Prevalence and Trends in Management of Idiopathic Normal Pressure Hydrocephalus in the United States: Insights from the National Inpatient Sample’. World Neurosurgery, 145, pp. e38–e52. doi: 10.1016/j.wneu.2020.09.012.

4. Andreão, F. F., Ferreira, M. Y., Brenner, L. B. O., Sousa, M. P., Palavani, L. B., Rairan, L. G., Tinti, I. S. U., Júnyor, F. D. S., Batista, S., Bertani, R., Amarillo, D. G. and Daccach, F. H. (2024). ‘Effectiveness and Safety of Ventriculoperitoneal Shunt Versus Lumboperitoneal Shunt for Idiopathic Intracranial Hypertension: A Systematic Review and Comparative Meta-Analysis’. World Neurosurgery, 185, pp. 359–369.e2. doi: 10.1016/j.wneu.2024.02.095.

5. Azad, T. D., Zhang, Y., Varshneya, K., Veeravagu, A., Ratliff, J. K. and Li, G. (2020). ‘Lumboperitoneal and Ventriculoperitoneal Shunting for Idiopathic Intracranial Hypertension Demonstrate Comparable Failure and Complication Rates’. Neurosurgery, 86 (2), pp. 272–280. doi: 10.1093/neuros/nyz080.

6. Bergsneider, M., Black, P. McL., Klinge, P., Marmarou, A. and Relkin, N. (2005). ‘Surgical Management of Idiopathic Normal-pressure Hydrocephalus’. Neurosurgery, 57 (3), pp. S2–29-S2-39. doi: 10.1227/01.NEU.0000168186.45363.4D.

7. Delwel, E. J., De Jong, D. A., Dammers, R., Kurt, E., Van Den Brink, W. and Dirven, C. M. F. (2013). ‘A randomised trial of high and low pressure level settings on an adjustable ventriculoperitoneal shunt valve for idiopathic normal pressure hydrocephalus: results of the Dutch evaluation programme Strata shunt (DEPSS) trial’. *Journal of Neurology*, Neurosurgery & Psychiatry, 84 (7), pp. 813–817. doi: 10.1136/jnnp-2012-302935.

8. DerSimonian, R. and Kacker, R. (2007). ‘Random-effects model for meta-analysis of clinical trials: An update’. Contemporary Clinical Trials, 28 (2), pp. 105–114. doi: 10.1016/j.cct.2006.04.004.

9. Gallegos, M., Morgan, M. L., Cervigni, M., Martino, P., Murray, J., Calandra, M., Razumovskiy, A., Caycho-Rodríguez, T. and Gallegos, W. L. A. (2022). ‘45 Years of the mini-mental state examination (MMSE): A perspective from ibero-america’. Dementia & Neuropsychologia, 16 (4), pp. 384–387. doi: 10.1590/1980-5764-dn-2021-0097.

10. Giordan, E., Palandri, G., Lanzino, G., Murad, M. H. and Elder, B. D. (2019). ‘Outcomes and complications of different surgical treatments for idiopathic normal pressure hydrocephalus: a systematic review and meta-analysis’. Journal of Neurosurgery, 131 (4), pp. 1024–1036. doi: 10.3171/2018.5.JNS1875.

11. Graham, P., Howman-Giles, R., Johnston, I. and Besser, M. (1982). ‘Evaluation of CSF shunt patency by means of technetium-99m DTPA’. Journal of Neurosurgery, 57 (2), pp. 262–266. doi: 10.3171/jns.1982.57.2.0262.

12. GraphPad Software (2024). GraphPad Prism [Computer software]. Available at: https://www.graphpad.com/ (Accessed: 15 December 2024).

13. Ho, Y., Chiang, W., Huang, H., Lin, S. and Tsai, S. (2023). ‘Effectiveness and safety of ventriculoperitoneal shunt versus lumboperitoneal shunt for communicating hydrocephalus: A systematic review and meta-analysis with trial sequential analysis’. CNS Neuroscience & Therapeutics, 29 (3), pp. 804–815. doi: 10.1111/cns.14086.

14. Hu, Y., Cao, C., Li, M., He, H., Luo, L. and Guo, Y. (2024). ‘Association between idiopathic normal pressure hydrocephalus and Alzheimer’s disease: a bidirectional Mendelian randomization study’. Scientific Reports, 14 (1), p. 22744. doi: 10.1038/s41598-024-72559-w.

15. Hung, A. L., Moran, D., Vakili, S., Fialho, H., Sankey, E. W., Jusué-Torres, I., Elder, B. D., Goodwin, C. R., Lu, J., Robison, J. and Rigamonti, D. (2016). ‘Predictors of Ventriculoperitoneal Shunt Revision in Patients with Idiopathic Normal Pressure Hydrocephalus’. World Neurosurgery, 90, pp. 76–81. doi: 10.1016/j.wneu.2016.02.061.

16. Kobayashi, E., Kanno, S., Kawakami, N., Narita, W., Saito, M., Endo, K., Iwasaki, M., Kawaguchi, T., Yamada, S., Ishii, K., Kazui, H., Miyajima, M., Ishikawa, M., Mori, E., Tominaga, T., Tanaka, F. and Suzuki, K. (2022). ‘Risk factors for unfavourable outcomes after shunt surgery in patients with idiopathic normal-pressure hydrocephalus’. Scientific Reports, 12 (1), p. 13921. doi: 10.1038/s41598-022-18209-5.

17. Kubo, Y., Kazui, H., Yoshida, T., Kito, Y., Kimura, N., Tokunaga, H., Ogino, A., Miyake, H., Ishikawa, M. and Takeda, M. (2008). ‘Validation of Grading Scale for Evaluating Symptoms of Idiopathic Normal-Pressure Hydrocephalus’. Dementia and Geriatric Cognitive Disorders, 25 (1), pp. 37–45. doi: 10.1159/000111149.

18. Medtronic (2023). Lumboperitoneal shunt: surgery - what to expect. Available at: https://www.medtronic.com/uk-en/patients/treatments-therapies/shunt-lumboperitoneal/getting-a-device/surgery-what-to-expect.html (Accessed: 15 December 2024).

19. Miyake, H. (2016). ‘Shunt Devices for the Treatment of Adult Hydrocephalus: Recent Progress and Characteristics’. Neurologia medico-chirurgica, 56 (5), pp. 274–283. doi: 10.2176/nmc.ra.2015-0282.

20. Moran, D., Hung, A., Vakili, S., Fialho, H., Jeon, L., Sankey, E. W., Jusué-Torres, I., Lu, J., Goodwin, C. R., Elder, B. D. and Rigamonti, D. (2016). ‘Comparison of outcomes between patients with idiopathic normal pressure hydrocephalus who received a primary versus a salvage shunt’. Journal of Clinical Neuroscience, 29, pp. 117–120. doi: 10.1016/j.jocn.2015.12.009.

21. Nakajima, M., Yamada, S., Miyajima, M., Ishii, K., Kuriyama, N., Kazui, H., Kanemoto, H., Suehiro, T., Yoshiyama, K., Kameda, M., Kajimoto, Y., Mase, M., Murai, H., Kita, D., Kimura, T., Samejima, N., Tokuda, T., Kaijima, M., Akiba, C., Kawamura, K., Atsuchi, M., Hirata, Y., Matsumae, M., Sasaki, M., Yamashita, F., Aoki, S., Irie, R., Miyake, H., Kato, T., Mori, E., Ishikawa, M., Date, I., Arai, H., and The research committee of idiopathic normal pressure hydrocephalus. (2021). ‘Guidelines for Management of Idiopathic Normal Pressure Hydrocephalus (Third Edition): Endorsed by the Japanese Society of Normal Pressure Hydrocephalus’. Neurologia medico-chirurgica, 61 (2), pp. 63–97. doi: 10.2176/nmc.st.2020-0292.

22. Page, M. J., McKenzie, J. E., Bossuyt, P. M., Boutron, I., Hoffmann, T. C., Mulrow, C. D., Shamseer, L., Tetzlaff, J. M., Akl, E. A., Brennan, S. E., Chou, R., Glanville, J., Grimshaw, J. M., Hróbjartsson, A., Lalu, M. M., Li, T., Loder, E. W., Mayo-Wilson, E., McDonald, S., McGuinness, L. A., Stewart, L. A., Thomas, J., Tricco, A. C., Welch, V. A., Whiting, P. and Moher, D. (2021). ‘The PRISMA 2020 statement: an updated guideline for reporting systematic reviews’. BMJ, p. n71. doi: 10.1136/bmj.n71.

23. Rinaldo, L., Bhargav, A. G., Nesvick, C. L., Lanzino, G. and Elder, B. D. (2020). ‘Effect of fixed-setting versus programmable valve on incidence of shunt revision after ventricular shunting for idiopathic normal pressure hydrocephalus’. Journal of Neurosurgery, 133 (2), pp. 564–572. doi: 10.3171/2019.3.JNS183077.

24. Ringstad, G., Vatnehol, S. A. S. and Eide, P. K. (2017). ‘Glymphatic MRI in idiopathic normal pressure hydrocephalus’. Brain, 140 (10), pp. 2691–2705. doi: 10.1093/brain/awx191.

25. Rostgaard, N., Olsen, M. H., Ottenheijm, M., Drici, L., Simonsen, A. H., Plomgaard, P., Gredal, H., Poulsen, H. H., Zetterberg, H., Blennow, K., Hasselbalch, S. G., MacAulay, N. and Juhler, M. (2023). ‘Differential proteomic profile of lumbar and ventricular cerebrospinal fluid’. Fluids and Barriers of the CNS, 20 (1), p. 6. doi: 10.1186/s12987-022-00405-0.

26. Sahuquillo, J., Rosas, K., Calvo, H., Alcina, A., Gándara, D., López-Bermeo, D. and Poca, M.-A. (2021). ‘How to Choose a Shunt for Patients with Normal Pressure Hydrocephalus: A Short Guide to Selecting the Best Shunt Assembly’. Journal of Clinical Medicine, 10 (6), p. 1210. doi: 10.3390/jcm10061210.

27. Scholz, R., Lemcke, J., Meier, U. and Stengel, D. (2018). ‘Efficacy and safety of programmable compared with fixed anti-siphon devices for treating idiopathic normal-pressure hydrocephalus (iNPH) in adults – SYGRAVA: study protocol for a randomized trial’. Trials, 19 (1), p. 566. doi: 10.1186/s13063-018-2951-6.

28. Serarslan, Y., Yilmaz, A., Çakır, M., Güzel, E., Akakin, A., Güzel, A., Urfalı, B., Aras, M., Kaya, M. E. and Yılmaz, N. (2017). ‘Use of programmable versus nonprogrammable shunts in the management of normal pressure hydrocephalus: A multicenter retrospective study with cost–benefit analysis in Turkey’. Medicine, 96 (39), p. e8185. doi: 10.1097/MD.0000000000008185.

29. Subramanian, H. E., Mahajan, A., Sommaruga, S., Falcone, G. J., Kahle, K. T. and Matouk, C. C. (2018). ‘The Subjective Experience of Patients Undergoing Shunt Surgery for Idiopathic Normal Pressure Hydrocephalus’. World Neurosurgery, 119, pp. e46–e52. doi: 10.1016/j.wneu.2018.06.209.

30. Turner, M. S. (1995). ‘The treatment of hydrocephalus: a brief guide to shunt selection’. Surgical Neurology, 43 (4), pp. 314–319; discussion 319-323. doi: 10.1016/0090-3019(95)80056-m.

31. Van Swieten, J. C., Koudstaal, P. J., Visser, M. C., Schouten, H. J. and Van Gijn, J. (1988). ‘Interobserver agreement for the assessment of handicap in stroke patients.’ Stroke, 19 (5), pp. 604–607. doi: 10.1161/01.STR.19.5.604.

32. Viechtbauer, W. (2010). ‘Conducting Meta-Analyses in *R* with the **metafor** Package’. Journal of Statistical Software, 36 (3). doi: 10.18637/jss.v036.i03.

33. Williams, M. A. and Malm, J. (2016). ‘Diagnosis and Treatment of Idiopathic Normal Pressure Hydrocephalus’. Continuum, 22 (2), pp. 579–599. doi: 10.1212/CON.0000000000000305.

34. Xiao, H., Hu, F., Ding, J. and Ye, Z. (2022). ‘Cognitive Impairment in Idiopathic Normal Pressure Hydrocephalus’. Neuroscience Bulletin, 38 (9), pp. 1085–1096. doi: 10.1007/s12264-022-00873-2.

35. Xie, D., Chen, H., Guo, X. and Liu, Y. (2021). ‘Comparative study of lumboperitoneal shunt and ventriculoperitoneal shunt in the treatment of idiopathic normal pressure hydrocephalus’. American Journal of Translational Research, 13 (10), pp. 11917–11924.

36. Yamada, S., Ishikawa, M., Nakajima, M. and Nozaki, K. (2022). ‘Reconsidering Ventriculoperitoneal Shunt Surgery and Postoperative Shunt Valve Pressure Adjustment: Our Approaches Learned From Past Challenges and Failures’. Frontiers in Neurology, 12, p. 798488. doi: 10.3389/fneur.2021.798488.

